# A non-competing pair of human neutralizing antibodies block COVID-19 virus binding to its receptor ACE2

**DOI:** 10.1101/2020.05.01.20077743

**Authors:** Yan Wu, Feiran Wang, Chenguang Shen, Weiyu Peng, Delin Li, Cheng Zhao, Zhaohui Li, Shihua Li, Yuhai Bi, Yang Yang, Yuhuan Gong, Haixia Xiao, Zheng Fan, Shuguang Tan, Guizhen Wu, Wenjie Tan, Xuancheng Lu, Changfa Fan, Qihui Wang, Yingxia Liu, Chen Zhang, Jianxun Qi, George Fu Gao, Feng Gao, Lei Liu

**Author notes:** These authors contributed equally (Yan Wu, Feiran Wang, Chenguang Shen, Weiyu Peng, Delin Li). Correspondence (Y.W.); (F.G.), (G.F.G.), (L.L.).

## Abstract

Neutralizing antibodies could be antivirals against COVID-19 pandemics. Here, we report the isolation of four human-origin monoclonal antibodies from a convalescent patient in China. All of these isolated antibodies display neutralization abilities in vitro. Two of them (B38 and H4) block the binding between RBD and vial cellular receptor ACE2. Further competition assay indicates that B38 and H4 recognize different epitopes on the RBD, which is ideal for a virus-targeting mAb-pair to avoid immune escape in the future clinical applications. Moreover, therapeutic study on the mouse model validated that these two antibodies can reduce virus titers in the infected mouse lungs. Structure of RBD-B38 complex revealed that most residues on the epitope are overlapped with the RBD-ACE2 binding interface, which explained the blocking efficacy and neutralizing capacity. Our results highlight the promise of antibody-based therapeutics and provide the structural basis of rational vaccine design.

**One Sentence Summary:** A pair of human neutralizing monoclonal antibodies against COVID-19 compete cellular receptor binding but with different epitopes, and with post-exposure viral load reduction activity.

## Main Text

The COVID-19 caused by the novel coronavirus COVID-19 virus has become a global health crisis. The virus has spread worldwide, causing fever, severe respiratory illness and pneumonia(*1, 2*). Phylogenetic analysis indicates that the emerging pathogen is closely related to several bat coronaviruses and to severe acute respiratory syndrome coronavirus (SARS-CoV)(*3-5*). It is worth to note that COVID-19 virus appears to be more easily transmitted from person to person(*6*). To date, however, no specific drugs or vaccines are available so far, except for several general antiviral drugs (eg. Remdesivir and hydroxychloroquine, et al.) under clinical investigation.

COVID-19 virus belongs to betacoronavirus genus, in which five human infected pathogens are implicated(*7, 8*). Among them, SARS-CoV and Middle East respiratory syndrome coronavirus (MERS-CoV) are two highly pathogenic viruses. As with other coronaviruses, the spike glycoprotein (S) homotrimer on the COVID-19 virus surface plays an essential role in receptor binding, triggering the cell membrane fusion for virus entry. The S protein is a class I fusion protein and each S protomer consists of S1 and S2 domains(*9*). The receptor binding domain (RBD) locates on the S1 region(*8*). Previous studies revealed that COVID-19 virus infects host cells by using the same receptor ACE2 as SARS-CoV for entry(*3, 10-12*), and the RBD undergoes hinge-like conformational movements to engage ACE2 binding(*13*). Numerous neutralizing antibodies targeting RBD of either SARS-CoV or MERS have been identified(*14-16*). Therefore, screening the neutralizing antibodies against COVID-19 virus by using the RBD protein is the priority strategy.

We expressed COVID-19 virus RBD protein as a bait to isolate the specific single memory B-cells from peripheral blood mononuclear cells (PBMCs) of a convalescent COVID-19 patient. The variable regions encoding the heavy chain and light chain were amplified from single B cells, respectively, and were then cloned into pCAGGS vector with the constant region to produce IgG1 antibodies as previous described(*17*). 17 paired B cell clones were amplified and 3 of which were identical (B5, B59 and H1). To identify the binding abilities of these antibodies, the plasmids containing the paired heavy chain and light chains were co-transfected into HEK 293T cells for monoclonal antibodies (mAbs) production. The supernatants were then screened for binding to COVID-19 virus RBD by Bio-Layer Interferometry (BLI) in OctetRED96, and an irrelevant anti-SFTSV Gn antibody and a SARS-CoV specific antibody were used as controls. The supernatants from four different antibodies (B5, B38, H2 and H4) exhibit the binding abilities to COVID-19 virus RBD, but failed binding to SARS-CoV RBD (Figure S1), suggesting that the epitope between SARS-CoV and COVID-19 virus RBDs are immunologically distinct. The usage of both heavy chain (V_H_) and light chain (V_L_) variable genes are listed in Table S1.

We then further measured the binding kinetics using surface plasmon resonance (SPR). These four antibodies displayed various binding abilities to COVID-19 virus RBD, with K_d_ ranging from 10^-7^ to 10^-9^ M (Figure1 A-D). H4 presented relatively higher binding affinity, with K_d_ of 4.48 nM, while B5 displayed relatively weaker binding affinity, with K_d_ of 305 nM. B38 and H2 bind to RBD, with K_d_ of 70.1 nM and 14.3 nM, respectively. We next studied the neutralizing activities against COVID-19 virus virus (BetaCoV/Shenzhen/SZTH-003/2020). All these four antibodies exhibit neutralizing activities, with IC_50_ values ranging from 0.177 µg/ml to 1.375 µg/ml (Figure 2). B38 was the most potent antibody, followed by H4, H2 and B5. Combined with the results of the competitive binding assay, it is suggested that different mAbs may exert neutralizing activities through distinct mechanisms.

**Figure 1.**
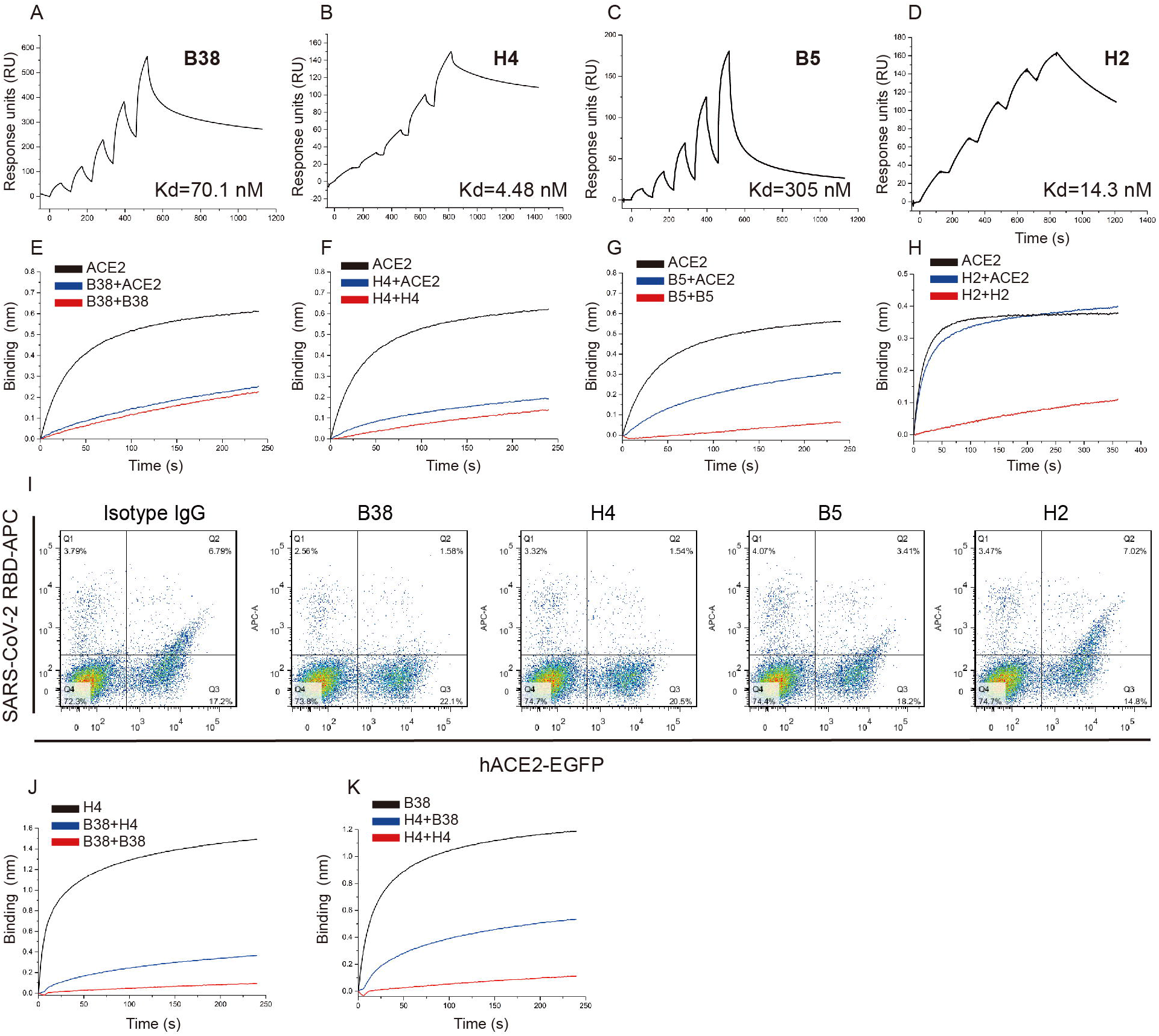
Characterization of COVID-19 virus specific neutralizing antibodies. (A-D) The binding kinetics of the four antibodies (B38, H4 B5 and H2) with COVID-19 virus RBD were measured by using a single-cycle BIAcore 8K system. The antibodies were captured on a CM5 chip coupled with anti-human Fc antibody. The COVID-19 RBD was serially diluted and injected with single-cycle kinetics procedure. The binding affinity (Kd) was labeled accordingly. (E-H) Competition binding to the COVID-19 virus RBD between antibody and ACE2 was measured by BLI in OctetRed96. Biotinylated COVID-19 virus RBD (10 µg/mL) were immobilized on SA sensors. The SA sensors were saturated with antibodies, and then flowed with corresponding antibody in the presence of 300 nM soluble ACE2 (blue) or without ACE2 (red). As a control, the immobilized biotinylated COVID-19 virus RBD was first flowed with buffer, and then flowed with the equal molar of ACE2 (black). (I) hACE2-EGFP was expressed on the HEK293T cell surface, and the cells were stained with 200 ng/mL COVID-19 virus RBD his-tag proteins pre-incubated with isotype IgG, B38, H4, B5 or H2. The percentage of both anti-histag APC^+^ and EGFP^+^ cells were calculated. (J-K) Competition binding to COVID-19 virus RBD between B38 and H4 was measured by BLI. Immobilized COVID-19 virus RBD (10 µg/mL) were saturated with 300 nM of the first antibody, and then flowed with equal molar of the first antibody in the presence of (blue) or without (red) the second antibody. As a control, the immobilized COVID-19 RBD was first flowed with buffer, and then flowed with the second antibody (black).

**Figure 2.**
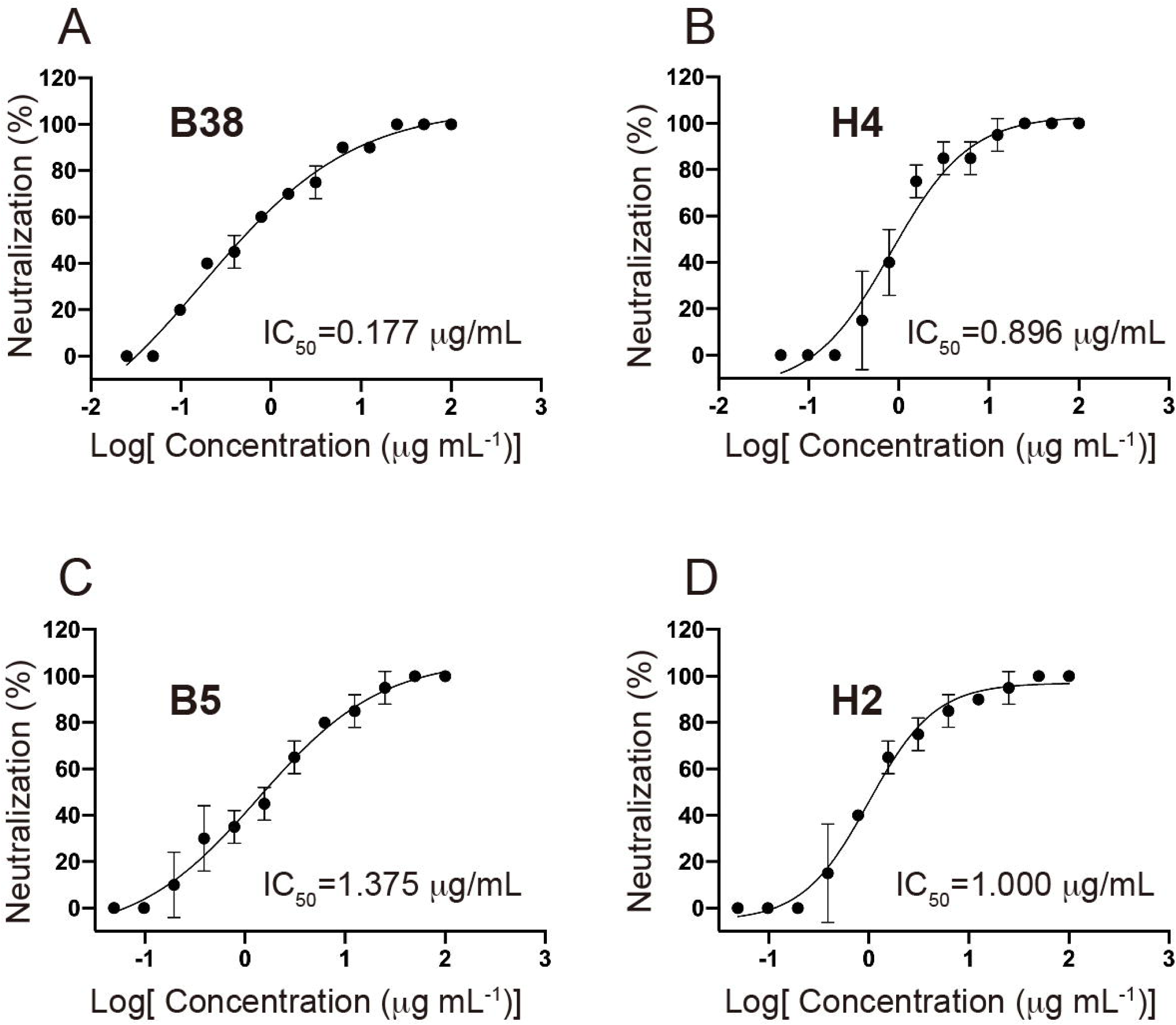
Four antibodies can effectively neutralize COVID-19 virus. The mixtures of SARS-CoV-2 and serially diluted antibodies were added to Vero E6 cells. After 5 days incubation, IC_50_ were calculated by fitting the CPE from serially diluted antibody to a sigmoidal dose-response curve. The IC_50_ were labeled accordingly.

To evaluate the competition abilities of each antibody for blocking the binding between viral RBD and ACE2, we performed the competition assay by using BLI in OctetRED96 and a blocking assay by using fluorescence-activated cell sorting (FACS). Specifically, the streptavidin biosensors labelled with biotinylated RBD were saturated with antibodies, and then flowed through with the test antibodies in the presence of soluble ACE2. B38 and H4 showed completely competition with ACE2 for the binding to RBD. In contrast, B5 displayed partial competition, while H2 show no competition with ACE2 for the RBD binding (Figure 1 E-H). The blocking assay by FACS presented similar results. Specifically, the ACE2-EGFP proteins were presented on the cell surface by transfection of ACE2-EGFP plasmid into HEK 293T cells, and then stained with RBD-histag proteins (positive control) or the preincubated RBD-antibody mixture. Both B38 and H4 can block the binding between RBD and ACE2 (Figure 1I), suggesting that the epitopes of these two antibodies locate on the RBD-ACE2 binding interface. To determine whether B38 and H4 target the same epitope, the epitope competition assay was performed with BLI. The viral RBD immobilized on the Ni-NTA sensor was first saturated with B38 IgG and flowed through B38 IgG in the presence or absence of H4 IgG, and vice versa. The result indicated that although RBD was saturated with the first antibody, the second antibody can still bind to RBD, suggesting that B38 and H4 recognize different epitopes on RBD with partial overlapping (Figure1 J-K).

To explore the protection efficacy of B38 and H4 against challenge with COVID-19 virus *in vivo*, therapeutic study was performed. hACE2 transgenic mice were administered with a single dose of 25 mg/kg of B38 or H4 12 hours after virus challenge. The results revealed that the body weight of B38 group substantially decreased and recovered at 3 days post infection (dpi) compared with PBS control group and H4 group (Figure 3A). The viral RNA copies of lung tissue were also detected at 3 dpi. The relative RNA copies change of both B38 group and H4 group were significantly lower than that of the PBS group, with a reduction of 32.8% and 26% RNA copies change of that in PBS group, respectively (Figure 3B). These results exhibit the identical trends to the neutralization abilities.

**Figure 3.**
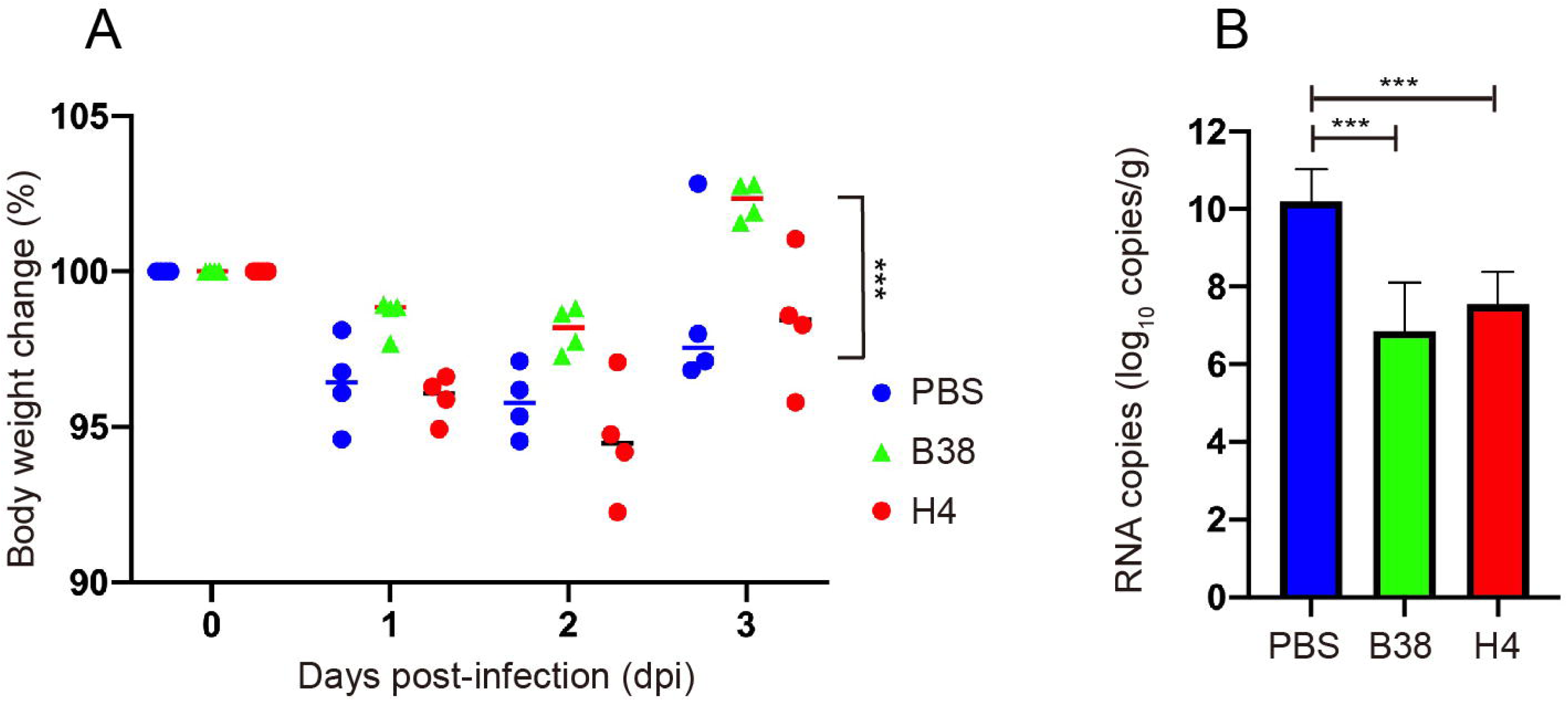
The protection efficiency of mAbs in hACE2 mice model post infection with COVID-19 virus. (A) Body weight loss were recorded for PBS (n=4), B38 treatment (n=4) and H4 treatment (n=4) groups. All the mice were challenged intranasally with COVID-19 virus, and 25 mg/kg antibodies were injected (i.p.) 12 hours post-infection. Equal volume of PBS was used as a control. The weight loss was recorded over three days, and the significant difference can be observed between B38 group and PBS group (unpaired *t*-test, \*\*\**p*<0.001). (B) The virus titer in lungs of three groups were determined at 3 dpi by qRT-PCR. The mAb treatment group can reduce the viral load in the lung of mice (unpaired *t*-test, \*\*\**p*<0.001).

To further elucidate the structural basis of the neutralization mechanisms of the antibodies, we prepared both the RBD-B38 and RBD-H4 Fab complexes by incubating the two proteins *in vitro* and then purified with a gel filtration column. Consistent with the binding affinity between viral RBD and B38 or H4, the stable complex was obtained (Figure S2). We therefore determined the crystal structure of B38 Fab with the viral RBD at 1.9 Å resolution (Table S2). Three complementarity-determining regions (CDRs) on the heavy chain (HCDR) and two CDRs on the light chain (LCDR) are involved in interaction with the viral RBD (Figure 4A and B). The buried surface area of heavy chain and light chain on the epitope is 713.9 Å and 497.7 Å, respectively. There are 36 residues in viral RBD involved in the interaction with B38, of which 21 residues and 15 residues interact with heavy chain and light chain, respectively (Table S3 and Figure 4B). Sequence alignment indicates that only 15 of the 36 residues in the epitope (defined as residues buried by B38) are conserved between COVID-19 virus and SARS-CoV (Figure 4D, 4F, and S3). This explains the specific reactivity of B38. Hydrophilic interactions are the major interacting forces on the interface between B38 and viral RBD (Table S4). Further analysis of the interactions of the binding interface reveals that the 470-loop of viral RBD plays an important role in binding to B38 HCDR1 loop (Figure 4G). The conformation of 470-loop is supported by Q474. In contrast, the corresponding loop (460-loop) in SARS-RBD displays distinct conformation due to the β-turn formed by P462 and G464, which is unfit for HCDR1 binding. S30 and S31 form hydrogen bonds to the side chain of K458, while the corresponding residue H445 in SARS-CoV RBD loses the interactions (Figure 4G). For the residues on contacting with the HCDR2, K417 is the only different residue between two RBDs involved in the interactions. The corresponding residue V404 in the SARS-CoV RBD loses the hydrogen bond to Y52, and the conformational change of the threonine results in losing the interaction with Y58 side chain (Figure 4H). Notably, the interactions between the side chains of two residues (Y100 and D103) on the HCDR3 and COVID-19 virus RBD are mediated by water molecules (Figure 4I). Specifically, the oxygen atom on the hydroxyl group interacts with E484, F490, L492 and Q493 on RBD via two water molecules. The side chain of D103 on the HCDR3 interacts with Y489 through a water molecule. Residue R97 on the HCDR3 hydrogen bonds to the side chains of both Y489 and N487. For the corresponding residues involved in the interactions with HCDR3 in SARS-CoV RBD, four residues are not conserved (N487, Y489, P491 and L492). P469 and W476 (corresponding residues F490 and E484 in COVID-19 virus RBD) are two unconserved residues between these two RBDs. E484 provides a polar contact with Y100 through water molecule, while P469 and W476 in SARS-CoV RBD are hydrophobic residues, which may be unsuitable for antibody binding. By contrast, residues involved in the interactions with LCDR loops between COVID-19 virus RBD and SARS-CoV RBD are more conserved (Figure 4J and K). Although two residues involved in the binding to LCDR3 loop are different between these two RBDs, they have the same properties (R403 and E406 in COVID-19 virus RBD, the corresponding residues are K491 and D493 in SARS-CoV RBD). This perfectly explains the B38 specific binding to COVID-19 rather than SARS-CoV To explore the structural basis for B38 blocking the interaction between COVID-19 virus RBD and ACE2, the complex structures of COVID-19 virus RBD/B38-Fab and COVID-19 virus RBD/hACE2 were superimposed. Both the V_H_ and V_L_ of B38 result in steric hindrance to the RBD binding to ACE2 (Figure 4C). Notably, the RBDs in B38-bound form and hACE2-bound form exhibit no significant conformational changes with the Cα root mean squared deviation (RMSD) of 0.489 Å (for 194 atoms). Further analysis indicated that 18 of the 21 amino acids on RBD are identical in binding B38 and ACE2 (Figure 4D), which clearly explains why B38 completely abolish the binding between COVID-19 virus RBD and the receptor.

**Figure 4.**
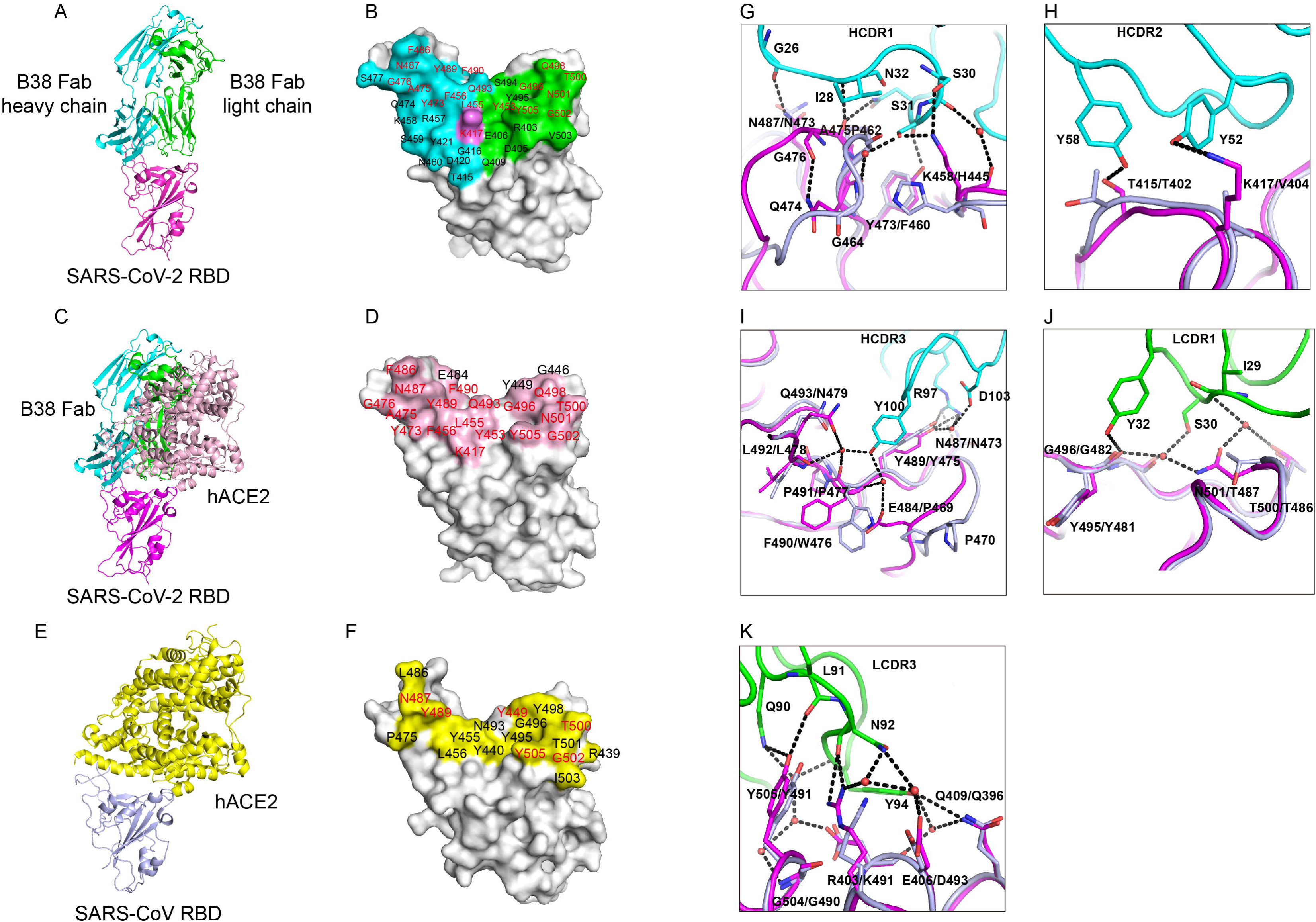
Structural analysis of B38 and COVID-19 virus RBD complex and the epitope comparison between B38 and hACE2. (A) The overall structure of B38 Fab and COVID-19 virus RBD complex. The B38 heavy chain (cyan), light chain (green) and COVID-19 virus RBD (magenta) are displayed in cartoon representation. (B) The epitope of B38 are shown in surface representation. The contacted residues by heavy chain, light chain or both are colored in cyan, green and magenta, respectively. The identical residues on RBD involved in binding to both B38 and hACE2 are labeled in red. (C) Superimposition of B38/COVID-19 virus RBD and hACE2/COVID-19 virus RBD (PDB: 6LZG). All the molecules are presented in cartoon, with the same colors in panel A. hACE2 is colored in light pink. (D) The residues involved in hACE2-RBD binding are highlighted in light pink. The identical residues on RBD involved in binding to both B38 and hACE2 are labeled in red. (E) The complex structure of SARS-CoV RBD (light blue) and hACE2 (yellow) (PDB:2AJF). (F) The residues of SARS-CoV RBD in contact with hACE2 are colored in yellow. The residues are numbered according to SARS-CoV RBD. The identical residues involved in hACE2 binding of two RBDs are labeled in red. (G-I) The detailed interactions between COVID-19 virus RBD and HCDR loops. (J-K) The detailed interactions between COVID-19 virus RBD and LCDR loops. The residues are shown in sticks with the identical colors to figure 3C. The water molecules are displayed in red sphere.

COVID-19 virus and SARS-CoV RBDs share relatively high identity, and both of them bind to receptor ACE2 for entry. However, the binding affinities are different. COVID-19 virus RBD exhibits higher binding affinity for receptor binding because of more atomic interactions with hACE2 than that of SARS-CoV RBD. Consequently, the different amino acids located on the hACE2 binding interface may lead to the generation of specific neutralizing antibodies in the host. So far, no antibody has been reported to cross-neutralize both SARS-CoV2 and SARS-CoV by competing the ACE2 binding site. CR3022 derived from a convalescent SARS patient can bind to RBDs from both viruses(18). However, it enables neutralizing SARS-CoV, but not COVID-19 virus. The epitope of CR3022 does not overlap with the ACE2-binding site(19), suggesting that it may be difficult to achieve cross-reactive neutralizing antibodies that can compete with ACE2 binding. A comprehensive understanding of the humoral immune responses against COVID-19 virus should be investigated in more patients. In addition, cocktail antibodies should be considered as an alternative therapeutic strategy to avoid potential escape mutants.

As the COVID-19 outbreak continues to spread, characterization of the epitopes on COVID-19 virus RBD protein is essential, which will provide valuable information to develop vaccines. Furthermore, the molecular features of the neutralizing antibodies targeting epitopes are helpful for the development of small molecule or peptide drugs/inhibitors. In conclusion, the neutralizing antibodies identified here are promising candidates for prophylactic and therapeutic treatment against COVID-19 virus.

## Data Availability

All the data and reagents presented in this study are available from the corresponding authors upon request.

## Materials and Methods

### Ethics statement

All procedures in this study involving authentic COVID-19 virus were performed in biosafety level 3 (BSL-3) facility. Murine studies were performed in an animal biosafety level 3 (ABSL-3) facility. All the procedures in the murine study were reviewed and approved by the Laboratory Animal Welfare and Ethics Committee in Chinese Academy of Sciences. The donor provided written informed consent for the use of blood and blood components followed the approval from the Research Ethics Committee of ShenZhen Third People’s Hospital, China.

### Cells and Viruses

HEK293T (ATCC CRL-3216) cells and Vero (ATCC CCL-81^TM^) cells were cultured at 37 °C with 5% CO_2_ in Dulbecco’s Modified Eagle medium (DMEM) supplemented with 10% fetal bovine serum (FBS). COVID-19 virus strain BetaCoV/shenzhen/SZTH-003/2020 (GISAID ID: EPI_ISL_406594) used in neutralization assay was isolated from the patient in Shenzhen Key Laboratory of Pathogen and Immunity, Shenzhen Third Peoples’ Hospital. COVID-19 virus strain HB-01 used in the animal experiment was provided by Professor Wenjie Tan from Chinese center for disease control and prevention. Vero E6 cells were applied to the amplification and titer titration of the virus stocks.

### Gene construction

The recombinant protein COVID-19 virus RBD was constructed into two vectors. Residues 331-532 (accession number EPI_ISL_402119) were cloned into pET21a for isolating mAbs as a bait. The coding sequences of COVID-19 RBD (residues 331-532), SARS-CoV RBD (residues 306-527, accession number NC_004718), and hACE2 (residues 19-615, accession number BAJ21180) were used in assays of SPR, BLI and crystal screening were constructed into pFastBac1 plasmid with an N-terminal gp67 signal peptide and a C-terminal six histidine tag. The pEGFP-N1-hACE2 plasmid was constructed by cloning the coding region into pEGFP-N1 vector using restriction enzymes Xhol and Smal.

### Protein expression and purification

The COVID-19 virus RBD recombinant protein was expressed in *E.coli* as inclusion body and in Bac-to-Bac expression system as soluble protein, respectively. The inclusion body was refolded and further purified by Superdex 200 Hiload 16/60 column (GE Healthcare) in 20 mM Tris, pH 9.0, 150 mM NaCl. Human ACE2 protein was expressed in baculovirus expression system. The constructed pFastBac1 plasmids were transformed into DH10Bac competent cells to generate recombinant bacmids. The bacmids were transfected into Sf9 cells to generate virus and then amplified the virus and infected High five cells for protein expression. Both the soluble COVID-19 virus RBD and hACE2 were purified by HisTrap HP column (GE Healthcare) and were further purified by size-exclusion chromatography with a Superdex 200 column (GE Healthcare) in 20 mM Tris, pH 8.0, 150 mM NaCl.

The plasmids of B38 heavy chain and light chain were co-transfected into HEK293T cells to produce B38 IgG. The full-length of B38 were used in neutralization and animal experiments. B38 Fab were generated by papain digestion and further purified by Protein A column (GE Healthcare) as described before(*20*).

### Bio-Layer Interferometry (BLI)

The antibody binding screening and the competitive binding of mAbs and hACE2 (or between two antibodies) were measured by BLI using the Octet RED96 system (FortéBio). All experiments were performed at 25 °C, and the biosensors were pre-equilibrated in a buffer containing 20 mM HEPES, pH7.4 and 150 mM NaCl and 0.005% (v/v) Tween-20 for 10min. For the antibody binding screening assay, 293T cell derived antibodies supernatants were loaded onto AHC biosensors for 120 s and flowed with 500 nM COVID-19 virus RBD or 1µM SARS-CoV RBD. To determine the competitive characteristics, 10 µg/mL biotinylated COVID-19 virus RBD was loaded onto streptavidin biosensors for 60 s, and flowed 300 nM of the first protein (one antibody) for 240 s and the second protein (hACE2 or the second antibody) for 240 s. The interference patterns from the biotinylated RBD with buffer and the uncoated biosensors with protein were analyzed as two sets of controls. We used corrected data to compare the competitive characteristics by Octet data analysis software.

### Surface plasmon resonance (SPR)

The affinity between COVID-19 virus RBD and antibodies were measured at room temperature using a BIAcore 8K system. All proteins used for kinetic analysis were exchanged to the buffer of 20 mM HEPES, 150 mM NaCl, pH 7.4 and 0.005% (v/v) Tween-20. A CM5 chip (GE Healthcare) was coupled with anti-human Fc antibody to capture the antibodies at 8000 response units. Gradient concentrations of COVID-19 virus RBD (from 200 nM to 12.5 nM with 2-fold dilution) were then flowed over the chip surface. After each cycle, the sensor surface was regenerated with Gly-HCl pH1.7. The affinity was calculated with BIAevaluation software.

### Crystal screening and structure determination

The COVID-19 virus RBD protein and B38-Fab fragment were mixed at a molar ration of 1:1.2. The mixture was incubated on ice for 60 min and further purified by Superdex-200 column (GE Healthcare). 5 mg/mL and 10 mg/mL of hCoV-19 RBD/B38 Fab proteins were used for crystal screening by vapor-diffusion sitting-drop method at 18°C. Diffracting crystals were obtained in the condition consisting of 0.15 M ammonium sulfate, 0.1 M MES pH 6 and 15% w/v PEG 400. Diffraction data were collected at Shanghai Synchrotron Radiation Facility (SSRF) BL17U (wavelength, 0.97919 Å). The crystals were cryo-protected by briefly soaking in reservoir solution supplemented with 20% (v/v) glycerol before flash-cooling in liquid nitrogen. The dataset was processed with HKL2000 software(*21*). The complex structure was determined by the molecular replacement method using Phaser with our previously reported hCoV-2 RBD structure (PDB code, 6LZG) and Fab structure (PDB code, 4TSA). The atomic models were completed with Coot(*22*) and refined with phenix.refine in Phenix(*23*), and the stereochemical qualities of the final models were assessed with MolProbity(*24*). Data collection, processing, and refinement statistics are summarized in Table S1. All the figures were prepared with Pymol software (http://www.pymol.org).

### Neutralization assay

10^4^ Vero cells were seeded in a 96-well plate (Costar) per well 24 hours before infection. On the day of infection, the cells were washed twice with PBS. Each mAb was diluted 2-fold in cell culture medium (modified eagle medium). 40 µL of diluted mAb (100 µg/mL as initial concentration) was added to 40 µL of cell culture medium containing 100 times the tissue culture infective dose (TCID_50_) of the BetaCoV/Shenzhen/SZTH-003/2020 strain virus on a 96-well plate in decuplicate and incubated at 37 °C for 2 hours. The mixture was then added to cells and incubated at 37 °C. The cytopathic effect was examined for 5 days post-infection. The complete absence of cytopathic effect in an individual culture well was defined as protection. The values of IC_50_ were calculated using prism software (GraphPad).

### Animal experiments

Twelve female hACE2 transgenic mice (5-6 weeks old) were divided into three groups with four mice in each group. All the mice were anaesthetized with thibromoethanol and then intranasally inoculated with 50µL 1×10^5^ TCID_50_ COVID-19 virus (HB-01). After 12 hours, mice received a dose of 25 mg/kg B38 or H4 i.p. in a volume of 100 µL, an equivalent volume of PBS was administered as a control. Body weights were monitored and recorded for four days. All the mice were euthanized at 3 dpi. The lung homogenates were prepared in 1 mL DMEM and then centrifuged at 3000 rpm for 10 min at 4 °C. The supernatant was collected to extract viral RNA by using the QIAamp Viral RNA Mini Kit (Qiagen). RNA was eluted in 80 µL elution buffer, and 5 µL was taken as the template for the quantitative real-time reverse transcription-PCR (qRT-PCR) by rRT-PCR kit (Mabsky Biotech Co., Ltd., CONFORMITE EUROPEENNE NO. DE/CA20/IVD-luxuslebenswelt-68/20). The primers targeting N gene were used as followed: F: 5’-ATTGGCATGGAAGTCACACCTTC-3’, R: 5’-TGCTTATTCAGCAAAATGACTTGAT-3’, probe: FAM--TGGTTGACCTACACAGGTGCCATCA--BHQ1. The amplification was performed as followed steps: 50 °C for 30 min, 95 °C for 3 min, followed by 45 cycles at 95 °C 5 s and 55 °C for 30 s. This mice model was established and supplied by Institute for Laboratory Animal Resources, NIFDC.

## Acknowledgements

We thank the staff of the BL17U1 beamline at Shanghai Synchrotron Radiation Facility (SSRF) for data collection. This work was supported by Zhejiang University special scientific research fund for COVID-19 prevention and control (2020XGZX019), the National Science and Technology Major Project (2018ZX10733403), the National Key R&D Program of China (2018YFC1200603), the National Key Plan for Scientific Research and Development of China (2016YFD0500304), National Natural Science Foundation of China (31872745 and 81902058), and the National Science and Technology Major Projects of Infectious Disease Funds (2017ZX10304402).

## Author contributions

Y.W., F.G., G.F.G. and L.L. initiated and coordinated the project. Y.W., F.G. and G.F.G designed the experiments. Y.L. and L.L. provided the convalescent PBMCs. Y.W. and C.S performed the cell sorting. W.P., C.Z, Z.L. and C.G.S. sequenced and constructed the antibodies. Y.W. conducted the SPR and Octet analysis with the help of F.Z., F.W., Z.L. and Q.W.. C.S. and Y.Y. evaluated the neutralizing potency. S.L. and Y.B. conducted the animal experiments with the help from G.W., W.T., X.L. C.F. and H.X.. qRT-PCR was conducted by Y.G.. F.W., W.P. D.L. C.Z. and Z.L expressed and purified proteins. J.Q. and F.G. collected the diffraction dada and determined the complex structure. Y.W., F.G. and G.F.G. analyzed the data and wrote the manuscript. S.T. revised the manuscript.

## References and notes

1. N. Zhu et al., A Novel Coronavirus from Patients with Pneumonia in China, 2019. N Engl J Med, (2020).

2. C. Wang, P. W. Horby, F. G. Hayden, G. F. Gao, A novel coronavirus outbreak of global health concern. Lancet 395, 470–473 (2020).

3. P. Zhou, Yang, X.L., Hu, B., Zhang, L., Zhang, W., Si, H.R., Zhu, Y., Li, B., Chen, J., Luo, H., Guo, H., Jiang, R.D., Liu, M.Q., Chen, Y., Shen, X.R., Wang, X., Zheng, X.S., Zhao, K., Chen, Q.J., Deng, F., Yan, B., Wang, Y.Y., Xiao, G.F., Shi, Z.L., A pneumonia outbreak associated with a new coronavirus of probable bat origin. Nature, (2020).

4. W. Tan et al., Notes from the Field: A novel coronavirus genome identified in a cluster of pneumonia cases-Wuhan, China 2019–2020. China CDC weekly 2, 2 (2020).

5. R. Lu et al., Genomic characterisation and epidemiology of 2019 novel coronavirus: implications for virus origins and receptor binding. Lancet 395, 565–574 (2020).

6. J. F. Chan et al., A familial cluster of pneumonia associated with the 2019 novel coronavirus indicating person-to-person transmission: a study of a family cluster. Lancet 395, 514–523 (2020).

7. F. Wu et al., A new coronavirus associated with human respiratory disease in China. Nature 579, 265–269 (2020).

8. G. Lu, Q. Wang, G. F. Gao, Bat-to-human: spike features determining ‘host jump’ of coronaviruses SARS-CoV, MERS-CoV, and beyond. Trends Microbiol 23, 468–478 (2015).

9. D. M. Knipe, P. M. Howley, Fields virology. (Wolters Kluwer/Lippincott Williams & Wilkins Health, Philadelphia, PA, ed. 6th, 2013), pp. 2 volumes.

10. A. C. Walls et al., Structure, Function, and Antigenicity of the SARS-CoV-2 Spike Glycoprotein. Cell, (2020).

11. M. Hoffmann et al., SARS-CoV-2 Cell Entry Depends on ACE2 and TMPRSS2 and Is Blocked by a Clinically Proven Protease Inhibitor. Cell, (2020).

12. Q. Wang et al., Structural and Functional Basis of SARS-CoV-2 Entry by Using Human ACE2. Cell, (2020).

13. D. Wrapp et al., Cryo-EM structure of the 2019-nCoV spike in the prefusion conformation. Science 367, 1260-1263 (2020).

14. Y. Zhou, Y. Yang, J. Huang, S. Jiang, L. Du, Advances in MERS-CoV Vaccines and Therapeutics Based on the Receptor-Binding Domain. Viruses 11, (2019).

15. L. Du et al., MERS-CoV spike protein: a key target for antivirals. Expert Opin Ther Targets 21, 131–143 (2017).

16. L. Du et al., The spike protein of SARS-CoV--a target for vaccine and therapeutic development. Nat Rev. Microbiol 7, 226–236 (2009).

17. Q. Wang et al., Neutralization mechanism of human monoclonal antibodies against Rift Valley fever virus. Nat Microbiol 4, 1231-1241 (2019).

18. X. Tian et al., Potent binding of 2019 novel coronavirus spike protein by a SARS coronavirus-specific human monoclonal antibody. Emerg Microbes & Infect 9, 382–385 (2020).

19. M. Yuan et al., A highly conserved cryptic epitope in the receptor-binding domains of SARS-CoV-2 and SARS-CoV Science, (2020).

20. Q. Wang et al., Neutralization mechanism of human monoclonal antibodies against Rift Valley fever virus. Nat Microbiol 4, 1231-1241 (2019).

21. Z. Otwinowski, Minor, W. & Charles W. Carter, Jr., Processing of X-ray diffraction data collected in oscillation mode. Methods Enzymol 276, 307–326 (1997).

22. P. Emsley, K. Cowtan, Coot: model-building tools for molecular graphics. Acta Crystallogr D Biol Crystallogr 60, 2126-2132 (2004).

23. P. D. Adams et al., PHENIX: building new software for automated crystallographic structure determination. Acta Crystallogr D Biol Crystallogr 58, 1948-1954 (2002).

24. C. J. Williams et al., MolProbity: More and better reference data for improved all-atom structure validation. Protein Sci 27, 293–315 (2018).

